# Pan T cells, Helper T cells, and Regulatory T cells are Associated with Negative Symptoms in Persons with Anti-Gliadin Antibody Positive Schizophrenia and Related Disorders

**DOI:** 10.1101/2025.02.24.25322815

**Authors:** Deepak Salem, Sarah M. Clark, Daniel J.O. Roche, Nevil J. Singh, Monica V. Talor, Robert W. Buchanan, Valerie Harrington, Zhenyao Ye, Shuo Chen, Deanna L. Kelly

## Abstract

**Background:** About one in three persons with a schizophrenia related disorder (SRD) have elevated anti-gliadin IgG antibodies (AGA). This AGA positive (AGA+) subgroup of SRD clinically has a higher burden of negative symptoms and are symptoms associated with high functional impairments with a lack of effective therapeutics. Alterations in T cells have been demonstrated in SRD, and we have previously shown regulatory T cells (Tregs) are increased and correlate with fewer negative symptoms in persons with SRD compared with healthy controls.

**Methods:** To further elucidate the role of the immune system in AGA+ SRD pathology, we investigated the relationship of T cells and negative symptoms in 26 medicated and clinically stable persons with SRD. Participants had blood drawn; AGA-IgG measured by ELISA (AGA positive defined as ≥20 U); had flow cytometry performed to quantify proportions of pan T cells (CD3+), helper T cells (CD3+CD4+), Tregs (CD3+CD4+CD25+Foxp3+), and activated Tregs (aTregs) (CD3+CD4+CD25+Foxp3+RA-); had serum cytokines measured; and completed the Scale for the Assessment of Negative Symptoms (SANS) to measure negative symptoms.

**Results:** 46% of persons with SRD in this study were AGA+ and, in this group specifically, pan-T cells were correlated with worse SANS total, anhedonia, alogia, and avolition (p<0.05), while helper T cells and Tregs were correlated with less negative symptoms (respectively, SANS total and alogia; SANS total, anhedonia, alogia; P<0.05). AGA+ persons with SRD also had several elevated serum cytokines, corresponding with a broadly pro-inflammatory phenotype.

**Conclusions:** These hypothesis-generating findings highlight T cell dysfunction in AGA+ positive SRD, suggesting Tregs protecting against negative symptom severity but also an unidentified other T cell population to possibly be driving negative symptom severity.

## 1. Introduction

Negative symptoms are enduring and prevalent in schizophrenia related disorders (SRD), contribute more to loss of function than positive symptoms, and may be the most significant predictor of poor long-term outcomes (Tamminga et al., 1998; Chang et al., 2016). Understanding negative symptom pathophysiology has been elusive, and as a result there are currently no Food and Drug Administration (FDA)-approved medications for negative symptoms associated with SRD.

Our group has previously identified and characterized a subgroup of people with SRD who show an immune and inflammatory response to gliadin, a protein found in wheat, barley and rye, and whose negative symptoms improve with a gluten free diet. Specifically, this subgroup, which represents about 33-38% of those with SRD, has high anti-gliadin antibodies (AGA-IgG) which correlate with peripheral inflammatory markers. Notably, this correlation, which has been replicated among several cohorts, is found only in the AGA-IgG positive (AGA+) but not AGA-IgG negative (AGA-) group. Additionally, this group displays attenuated positive symptom psychopathology (Cascella et al., 2011; Čiháková et al., 2018; Eaton et al., 2022; Jin et al., 2012; Kelly et al., 2018a; Lachance and McKenzie, 2014; Okusaga et al., 2013; Sidhom et al., 2012). Several lines of evidence indicate that the inflammatory response to gliadin may impact brain function, as indicated by peripheral AGA-IgG correlations to myoinositol and total choline (magnetic resonance spectroscopy; H^1^-MRS), high central AGA-IgG levels in cerebral spinal fluid, and the notable improvement in psychiatric symptoms, specifically negative symptoms (Cohen’s D=0.53), when gluten is removed from the diet (Rowland et al, 2017; Severence et al, 2015; Jackson et al, 2012; Kelly et al, 2019).

In a recent systematic review and meta-analysis it was found that negative symptoms are related to immune function in SRD (Dunleavy et al., 2022). In addition, several lines of evidence have pointed to immune cell dysfunction in SRD, including dysfunction in T cells – which are adaptive immune cells that mount potent cellular immune responses in an antigen-specific manner, by secreting cytokines or directly killing target cells (Ermakov et al., 2022). While such T cells can belong to either helper (T_H_) or cytotoxic (T_C_) lineages, a subset of T_H_ cells known are Regulatory T cells (Tregs) also function as potent immunosuppressive cells to reduce inflammation. Intriguingly, many markers of T cell activity including cytokines and inflammation are often altered in SRD (Pellicci et al., 2020).

Our work has shown that negative symptoms in SRD are negatively correlated to Tregs (Kelly et al., 2018c). We found the severity of alogia and affective blunting to decreased in those with more Tregs in SRD, possibly suggesting Tregs protecting emotional expression (Kelly et al., 2018c). We hypothesized then that Tregs may be dampening the inflammatory environment that was driving negative symptom pathology in SRD.

In other gluten related disorders, there is dysfunction in the immune system that results in a pro-inflammatory phenotype at both the gut and systemic levels (Leonard et al., 2017; Volta et al., 2019). In a similar manner, we have shown in AGA+ SRD that serum pro-inflammatory cytokines are increased compared with AGA-SRD, suggesting a similar immune profile to gluten related disorders in people without SRD (Kelly et al., 2018b). However, to date, we are not aware of any studies assessing T cells or their relationship to negative symptoms in AGA+ SRD.

To further classify this unique subgroup of AGA+ SRD, we investigated T cell subtype distributions and their relationship with negative symptoms. Given what is known thus far about immune dysfunction in AGA+ SRD, we hypothesize that T cell populations in SRD may differ with AGA status and that T cell populations may correlate differently with negative symptoms in AGA+ versus AGA-SRD. We also hypothesize that serum cytokines – consistent with our prior work – may be elevated in AGA+ SRD compared with AGA-SRD.

## 2. Methods

### 2.1 Participants

We performed a secondary data analysis of a previous study assessing pan-T cells, helper T cells, and Tregs in medicated persons with SRD and healthy controls. Participants were recruited from the Maryland Psychiatric Research Center, University of Maryland School of Medicine. Participants included were those who met DSM-5 criteria for schizophrenia or schizoaffective disorder and were required to be clinically stable on the same antipsychotic treatment for at least 30 days prior to the study. All participants were able to provide informed consent. For the secondary data analysis performed in this study, we only included persons with SRD who were subsequently sorted into two groups: AGA+ SRD or AGA-SRD. Exclusion criteria in all participants included any current active systemic infection with fever (T > 38 C), chronic infections, autoimmune conditions, pregnancy, co-occurring DSM-V diagnoses of alcohol or other substance use disorders at any time in the last 3 months, current gluten free diet or other immunologically altering medical conditions.

### 2.2 Study Methods

This cross-sectional study included a one-time visit where participants were evaluated for symptom assessments and had a blood draw. After signing informed consent, all participants had a medical and psychiatric history completed. Each participant was evaluated for negative symptom psychopathology and severity using the Scale for the Assessment of Negative Symptoms (SANS). The SANS total score was assessed by using the total score excluding the global items, inappropriate affect, poverty of content of speech, and attention items (Andreasen, 1982; Buchanan et al., 2007; Kelly et al., 2018c).

### 2.3 Laboratory Measures

Blood was collected between 0700 and 0800 from fasting participants and maintained at 2-5 C and processed within 2 hours of collection. Pertinently, we collected a complete blood count (CBC), isolated peripheral blood mononuclear cells (PBMCs), and performed flow cytometric analysis utilizing BDTM LSR II flow cytometry, (BD Biosciences, San Jose, California) which was conducted as per previous methods. Cells were labeled for CD3, CD4, CD25, and CD45RA prior to fixation and permeabilization to stain for FOXP3. We gated lymphocytes on forward scatter versus side scatter, followed by CD3 and CD4 positivity, followed by CD25 and Foxp3 positivity, and finally, CD45RA and Foxp3. We defined pan T cells (CD3+ T cells) as a percentage of lymphocytes, helper T cells (CD3+CD4+ T cells) as a percentage of pan T cells, Tregs (CD3+CD4+CD25+Foxp3+ T cells) as a percentage of Helper T cells, and aTregs (CD3+CD4+CD25+Foxp3+CD45RA-T cells) as a percentage of Tregs. A representative flow chart and gating strategy is depicted in Figure 1.

**Figure 1:**
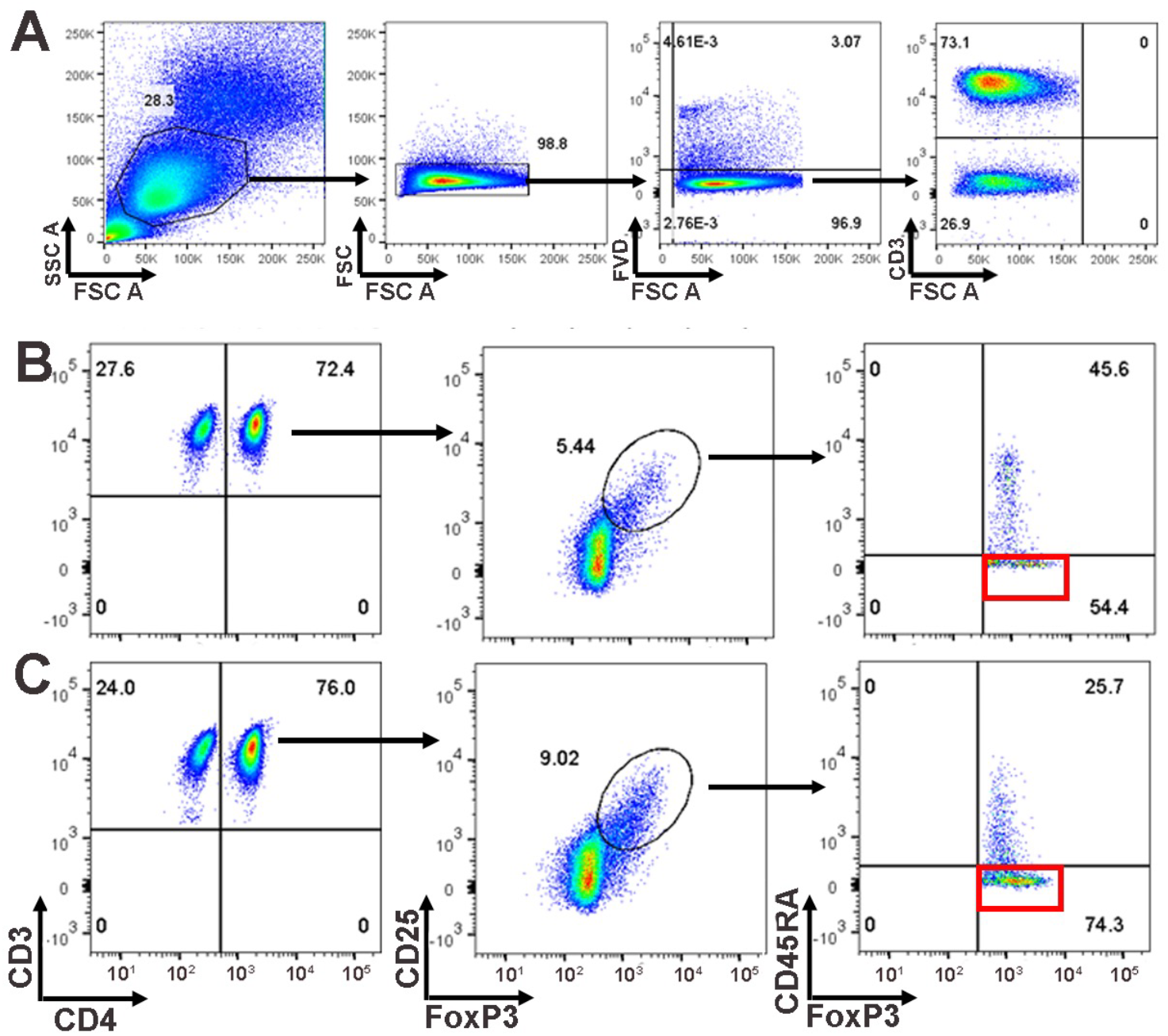
Gating strategy and flow charts. 1A) Representative gating strategy for CD3+ T cells. 1B-C) Representative flow charts for 1B) AGA+ SRD and 1E) AGA-SRD, illustrating the gating for the identification and quantification of activated Tregs (aTregs) (red rectangle).

We measured AGA IgG in the Čiháková Laboratory at Johns Hopkins University using the Werfen Diagnostics kit catalog # 708655. Units were determined by the manufacturer based on a semiquantitative detection of Gliadin IgG antibodies in human serum. AGA+ status was defined as ≥20 U AGA-IgG and AGA-status was defined as < 20 U AGA-IgG. We measured serum cytokine concentration by Luminex Multiplex at the Cytokine Core Laboratory at the University of Maryland School of Medicine, including the following cytokines with corresponding detection ranges: Tumor Necrosis Factor alpha (TNF-α; 7.2–1760□pg/mL), Interleukin-6 (IL-6; 3.2–790□pg/mL), Interleukin-10 (IL-10; 4.2–1030□pg/mL), Interleukin-2 (IL-2; 39– 9690□pg/mL), Interleukin-1 beta (IL-1β; 19–4800□pg/mL), Interferon gamma (IFN-*γ*; 47– 11460□pg/mL), Interleukin-4 (IL-4; 15–3690□pg/mL), Interleukin-17 (IL-17; 15–3740□pg/mL), C-C Motif Chemokine Ligand 28 (CCL28; 206–50060□pg/mL), Interleukin-13 (IL-13; 470– 114430□pg/mL), Transforming Growth Factor beta (TGF-β; 9.8–10000□pg/mL), Interleukin-35 (IL-35; 0.8–800□ng/mL), C-X-C Motif Chemokine Ligand 10 (CXCL10; 1.8–440□pg/mL), C-C Motif Chemokine Ligand 20 (CCL20; 8–1970□pg/mL), C-C Motif Chemokine Ligand 22 (CCL22; 49–11990□pg/mL), GALECTIN-1 (1417–344510□pg/mL), C-C Motif Chemokine Ligand 17 (CCL17; 96–23500□pg/mL), Interleukin-23 (IL-23; 160–39060□pg/mL), and Interleukin-18 (IL-18; 13–3380□pg/mL). All cytokines were log transformed prior to any analysis.

### 2.4 Statistical Analyses

All samples and patient information were de-identified for blinding. We reported demographic data utilizing descriptive statistics and when indicated, as mean ± SD. We tested demographic and clinical data between subgroups for possible confounders using unpaired two-tailed t-tests for continuous variables and utilized chi-square tests for categorical variables. Additionally, we utilized multiple linear regression in with inclusion of covariates of weight, gender, diagnosis, and smoking status to examine possible confounding effects. We again utilized multiple linear regression to assess for any impact of medications on outcomes. The above analyses were conducted in GraphPad PRISM (version 10.0.0 for Windows, GraphPad Software, Boston, Massachusetts USA, www.graphpad.com).

We utilized two tailed Mann-Whitney U to assess differences between T cell subtypes between AGA positive and negative SRD and we reported results as median and Interquartile Range (IQR). We utilized spearman’s correlation in determining the relationship between T cell subtypes and negative symptoms. We utilized a multiple interaction regression analysis to specifically look at the effect of AGA as a group variable affecting the interaction between T cells and negative symptoms, reported by β (the interaction coefficient) and 95% Confidence Interval (CI). Serum cytokine concentration was first sorted into different cytokine categories: Those that contained censored data (“non-detectable”, corresponding to being below the limit of detection) and those that did not. For cytokines with samples that did not have any non-detectable values (TGF-B, IL-35, CXCL10, CCL20, CCL22, GALECTIN-1, CCL17, IL-23 and IL-18), we utilized Mann-Whitney U to assess differences between AGA positive and AGA negative SRD and reported results as median and IQR. For cytokines with samples that had non-detectable values (TNF-A, IL-6, IL-10, IL-2, IL-1B, IFN-G, IL-4, IL-17, CCL28, IL-13), we converted individual numerical results into two categorical values: detectable (any value that is “non-detectable”, as in not below the limit of detection) and non-detectable. We then assessed for differences in the proportion of detectable values for each cytokine between AGA positive and negative SRD utilizing the fisher exact. Above analyses were conducted utilizing R version 4.3.3 (2024-02-29) and figures were made in RStudio using the ggplot2 package (Wickham, 2009). We did not correct for multiple comparisons due to the exploratory nature of this secondary data analysis. p < 0.05 is considered significant.

## 3. Results

### 3.1 Demographic Information

In total, 26 participants met eligibility criteria for having a diagnosis of SRD (mean age 46.0 ± 10.9; 73.1% male, 26.9% female). Prevalence of AGA-IgG positivity in SRD patients was 46.2% (12 of 26). In comparison of AGA status in SRD, there were no statistically significant differences in age, biological sex, race, weight, smoking status, white blood cell count, lymphocyte counts, medications – specifically in clozapine, first & second-generation antipsychotics, antidepressants, or anxiolytics (Table 1).

**Table 1:** T cells correlate with negative symptoms in positive but not negative or All AGA-IgG groups. Mann-Whitney U was utilized for all statistical comparisons. *p-value = < 0.05, **p-value < 0.01. Abbreviations: SRD = schizophrenia related disorders, rs = Spearman’s Correlation Coefficient

| Variable | SRD |  |  |
| --- | --- | --- | --- |
|  | Positive AGA-IgG (N=12) | Negative AGA-IgG (N=14) | Statistics |
| Age (years, mean ± SD) | 42.6 ± 10.9 | 48.1 ± 10.8 | p = 0.213 |
| Sex (male, N, %) | 10 M, 2 F | 9 M, 5 F | p = 0.27 |
| Race (N, %) | 4 White, 8 AA, 0 Asian, 0 Mixed | 5 White, 7 AA, 0 Asian, 2 Mixed | p = 0.56 |
| Cigarette smoker current | 4 yes, 8 no | 6 yes, 8 no | p = 0.62 |
| Weight (kg) | 99.6 ± 29.5 | 102.8 ± 38.3 | p = 0.811 |
| White blood cell count (x103/uL) | 6.0 ± 2.8 | 6.5 ± 2.9 | p = 0.63 |
| Lymphocytes (x103/uL) | 1.9 ± 0.9 | 1.7 ± 0.5 | p = 0.55 |
| Medications |  |  | p = 0.31 |
| Clozapine | N = 3 | N = 6 |  |
| Second Generation Antipsychotic | N = 2 | N = 5 |  |
| First Generation Antipsychotic | N = 7 | N = 3 |  |
| Antidepressants | N = 3 | N = 5 |  |
| Anxiolytics | N = 5 | N = 3 |  |

In addition, we performed multiple linear regression analysis with either Tregs, aTregs, helper T cells, or pan T cells as the dependent variable to assess the contribution of age, weight, and smoking status, which revealed these variables had no significant effect. We also performed this same analysis with the different T cell groups as the dependent variable to assess the effects of medications and found no statistically significant effects on any T cell group by first generation anti-psychotics, clozapine, other second-generation anti-psychotics, anti-depressants, or anti-anxiety medications.

### 3.2 AGA-IgG positivity stratifies the relationships between T cells and negative symptoms in SRD

We first sought to investigate whether there were differences in the proportion of pan T cells, helper T cells, Tregs, or aTregs when comparing AGA+ versus negative SRD. We found aTregs to be significantly increased in the AGA+ SRD group compared with AGA-IgG negative SRD (median=70.2 (21.9) vs 28.3 (15.5); p=0.0064) (Figure 2). There were no other statistically significant differences found between AGA positive and negative SRD in the proportions of pan T cells, helper T cells, or Tregs.

**Figure 2:**
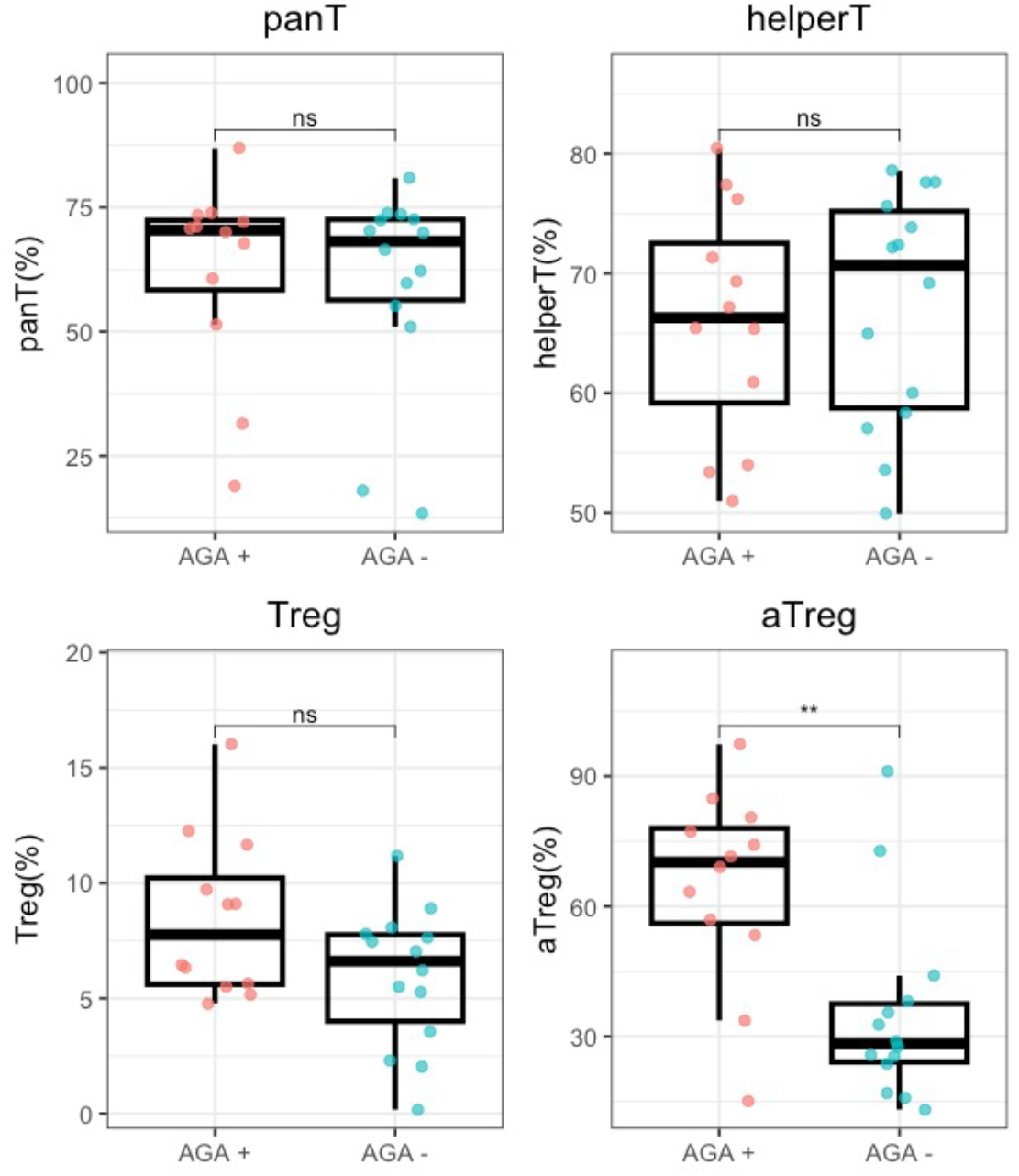
Average proportions of T cells differ in schizophrenia by AGA positivity. Values of each T cell population are graphed with each data point and box and whisker plots were generated in RStudio. Mann-Whitney U was utilized to determine statistical significance. The middle bar of each box plot shows the median value, bottom of the box showing the upper end of the lower quartile, the top of the box showing the lower end of the upper quartile, and the whiskers showing minimum and maximum values. Abbreviations: panT (pan T cell), defined as CD3+ T cells, helperT (helper T cell), defined as CD3+CD4+ T cells, Treg defined as CD3+CD4+CD25+Foxp3+ T cells, and aTregs defined as CD3+CD4+CD25+Foxp3+CD45RA-T cells. * indicates p < 0.05; ** indicates p < 0.01.

We next sought to investigate the relationship between T cells and negative symptoms in this AGA+ SRD population compared with AGA-IgG negative SRD (Figure 3). We notably found several associations that were statistically significant in the AGA positive SRD group. Pan T cells in AGA positive SRD positively correlated with SANS total (rs= 0.60, p=0.042), SANS anhedonia (rs = 0.63, p=0.032) SANS alogia (rs = 0.76, p=0.0061), and SANS avolition (rs=0.59, p= 0.046) (Figure S1A). In sharp contrast, we found helper T cells in AGA+ SRD to correlate negatively with both SANS total (rs= −0.62, p=0.034) and SANS alogia (rs = −0.77, p=0.0051) (Figure S1B. We also found Tregs in AGA+ SRD to negatively correlate with SANS total (rs = −0.64, p=0.031), SANS anhedonia (rs=-0.60, p=0.042), and SANS alogia (rs=-0.64, p=0.029) (Figure S1C). In this same AGA+ SRD group, there were no statistically significant correlations between any negative symptom category and aTregs (Figure S1D). Remarkably, we did not find any significant relationships between any T cell population and any negative symptom category in AGA-SRD.

**Figure 3:**
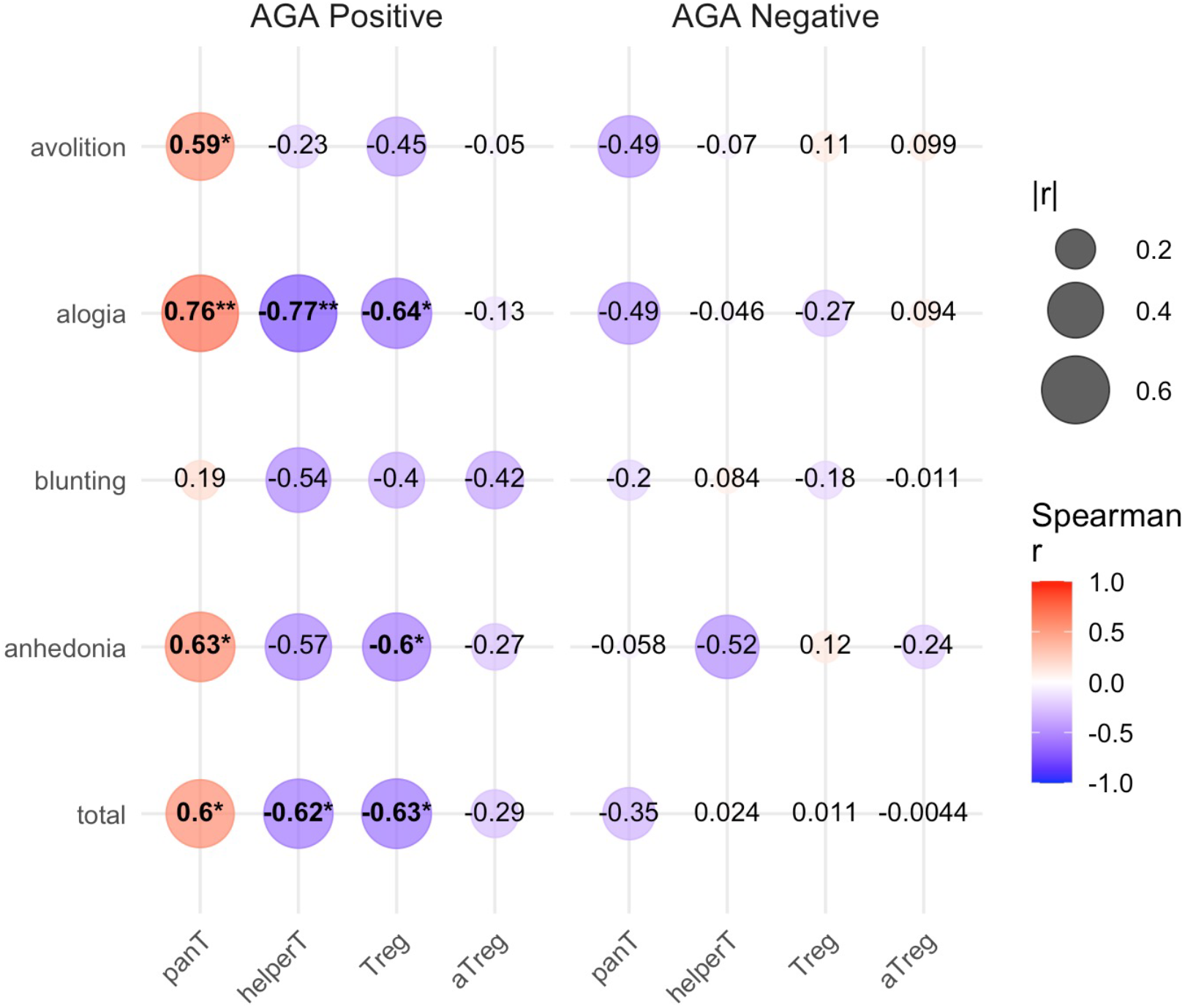
T cells correlate with negative symptoms in positive but not negative AGA-IgG schizophrenia. Spearman correlation coefficients (Spearman r, or “rs”) were calculated and heatmap was generated using RStudio. Bubble size correlates with the absolute value of rs and color correlates with positive or negative directionality (red is positive, blue is negative). Abbreviations: panT (pan T cell), defined as CD3+ T cells, helperT (helper T cell), defined as CD3+CD4+ T cells, Treg defined as CD3+CD4+CD25+Foxp3+ T cells, and aTregs defined as CD3+CD4+CD25+Foxp3+CD45RA-T cells. * indicates p < 0.05; ** indicates p < 0.01.

We also utilized multiple interaction regression to assess a mediating effect by AGA status itself on the relationship between T cell subtypes and negative symptoms. We found a significant interaction between AGA status and pan T cells on SANS Alogia (β = 0.825, 95% CI = [0.016, 1.63], p = 0.046). No other statistically significant interactions were found between AGA status on the relationship between T cell subtypes and negative symptoms.

### 3.3 Serum cytokines are increased in AGA positive SRD

To further characterize the immunologic landscape in AGA+ SRD, we investigated whether we could see differences in serum cytokine concentrations. In cytokines with continuous data, we found IL-35 (median= −0.13 (1.21) vs. median= −0.38 (0.61); p= 0.048) and CCL17 (median= 2.55 (0.18) vs. median= 2.37 (0.37); p= 0.037) to be significantly increased in AGA positive versus AGA negative SRD (Figure 4). Additionally, while they did not reach statistical significance, CCL20 (median= 2.77 (0.28) vs. median= 2.19 (0.11); p= 0.067) and IL-23 (median= 2.89 (0.34) vs. median= 2.24 (0.22); p= 0.060) were increased in AGA+ SRD compared with AGA-SRD and trended towards being significantly different. In cytokines with censored data, we found IL-1B (66.7% vs. 14.3%; p= 0.0093), IL-2 (58.3% vs. 7.1%; p= 0.0093), CCL28 (83.3% vs. 35.7%), and IL-13 (66.7% vs. 21.4%; p= 0.045) to have more detectable cytokines in AGA+ SRD versus AGA-SRD (Figure 5).

**Figure 4:**
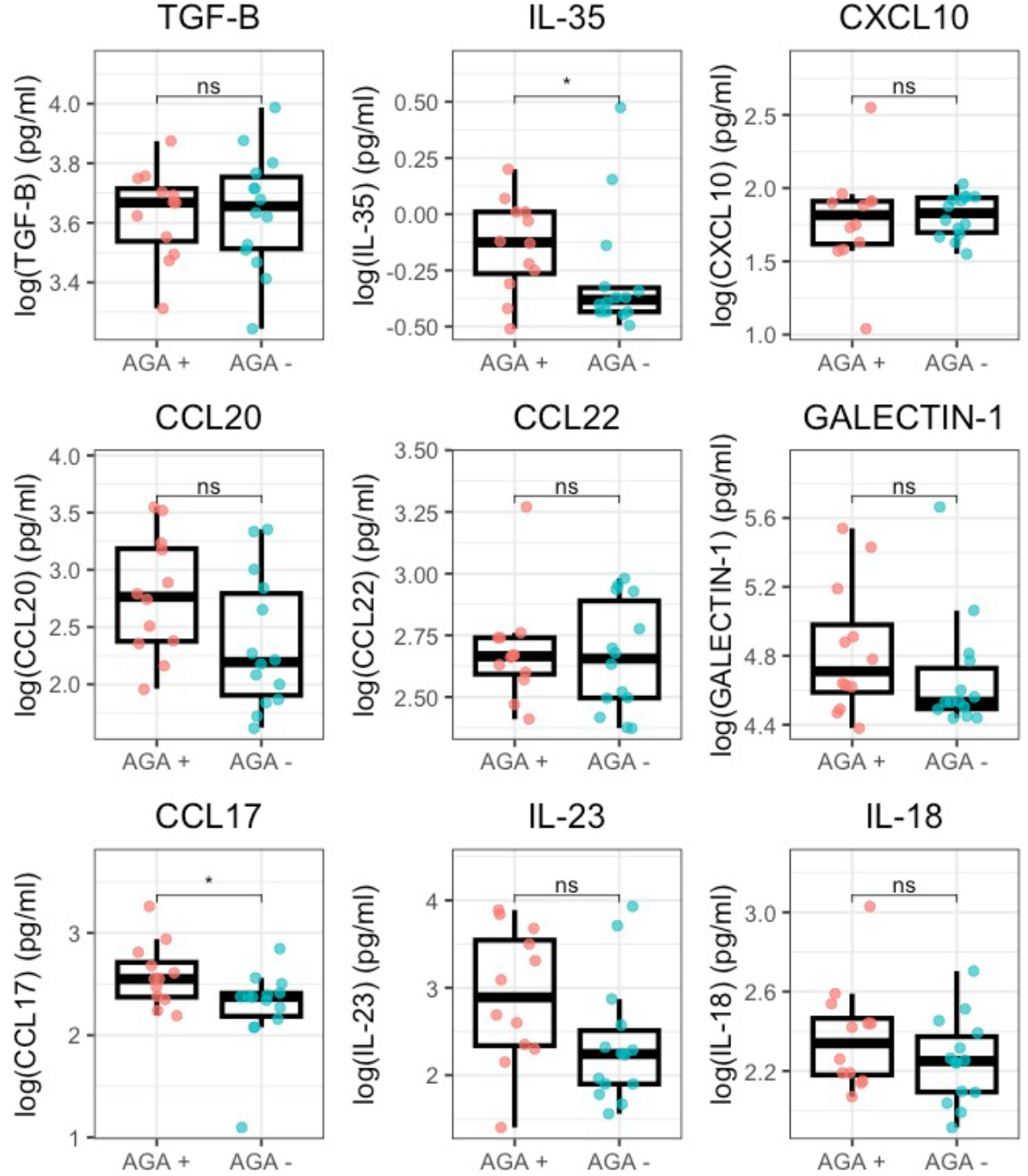
Cytokine concentrations differ in AGA positive vs AGA negative schizophrenia. Values of each cytokine were log10 transformed and then graphed with each data point visualized and box and whisker plots were generated in RStudio. The middle bar of each box plot shows the median value, bottom of the box showing the upper end of the lower quartile, the top of the box showing the lower end of the upper quartile, and the whiskers showing minimum and maximum values. * indicates p < 0.05; ** indicates p < 0.01.

**Figure 5:**
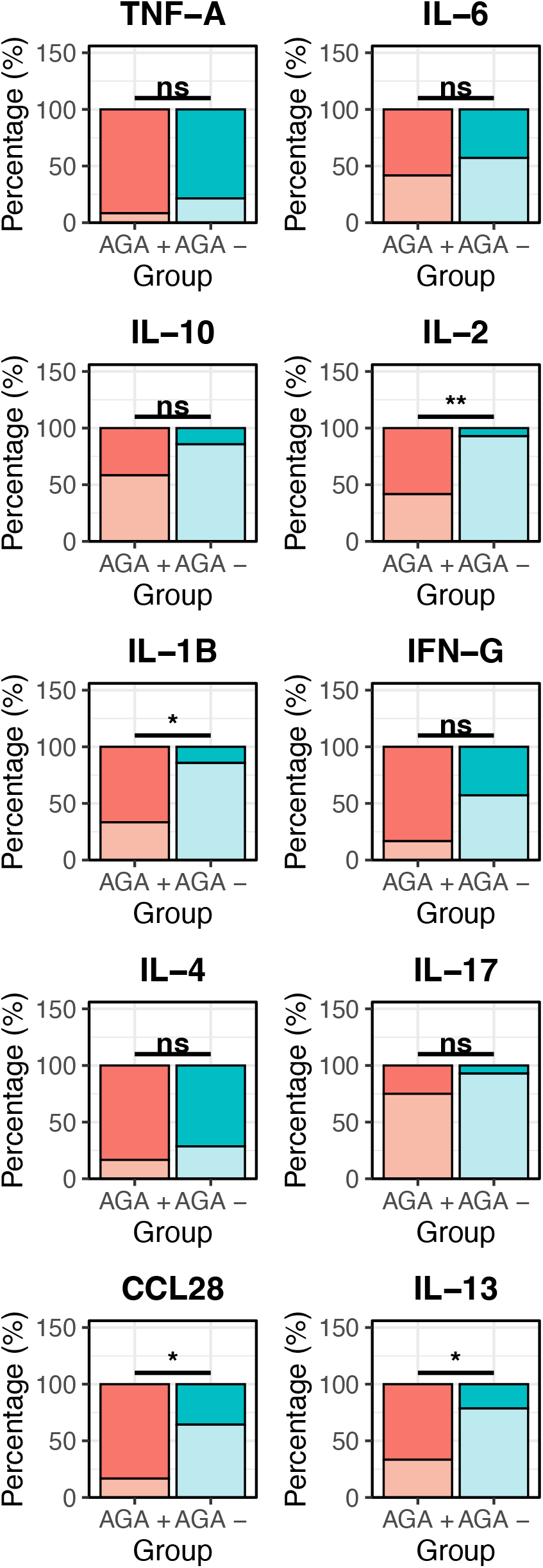
Cytokine concentrations differ in AGA positive vs AGA negative schizophrenia. Stacked bar plots show the amount of “Detectable (D)” values or “Non-Detectable (ND)” cytokine percentages for each cytokine between the two AGA groups. The darker shades for each color represent % D values and corresponding lighter shade are % ND values. Statistical significance was calculated using the Fisher Exact test to determine whether there is a significant difference in the amount of Detectable cytokine values in the two AGA groups. * indicates p < 0.05; ** indicates p < 0.01.

## 4. Discussion

To our knowledge, this is the first study assessing T cells in those with AGA-IgG positive titers in SRD. We found proportions of different T cell groups to be differentially associated with negative symptomatology in SRD depending on AGA status. In AGA+ but not AGA-SRD, we found pan T cells to be positively associated with negative symptoms; specifically, increased pan T cells were associated with worse emotional expression, worse speech production, and worse initiation and motivation in goal-directed behaviors. Conversely, we found both helper T cells and Tregs to be negatively associated with negative symptoms. In AGA+ but not AGA-SRD, increased helper T cells were associated with better speech production, and increased Tregs were associated with both better speech production and emotional expression. This sharp contrast in negative symptom correlations between pan-T cells and the helper T Cell and Treg lineages is notable, as helper T cells and Treg lineages do not make up the entirety of every cell population under the umbrella of “pan-T cells”, defined in this study as lymphocytes that are CD3 positive. From this data, it appears that cytotoxic CD8+ or unconventional T cells, that were not assayed in this study may be responsible for the strong positive correlation with negative symptoms in AGA+ SRD. This effect does not appear to be dependent on the proportion of cells and may represent a functional difference in these different T cell populations in AGA+ SRD, as total lymphocytes, pan-T cells, helper T cells, and Tregs were all statistically similar in proportions between AGA+ and negative SRD. Other T cell groups include Killer CD8+ T cells, natural Killer T cells (NKT cells), Gamma-Delta T cells (*γδ*-T cells), and mucosal-associated invariant T (MAIT) cells – which have shown dysfunction in SRD – and again may be contributing to the phenotype of AGA positive SRD (Müller et al., 1998; Pellicci et al., 2020; Sperner-Unterweger et al., 1999; Varun et al., 2019; Wang et al., 2023).

The differences in serum cytokine concentrations and correlations with T cell subtypes in AGA+ SRD continue to implicate a disrupted immune system in this sub-group of patients. Our finding of IL-1B (broadly pro-inflammatory) and IL-2 (important for proliferation of all T cells) being elevated in AGA SRD supports a generalized pro-inflammatory phenotype (Akkouh et al., 2020; Malek, 2008, p. 1). We also found IL-13, CCL17, and CCL28 to be elevated in AGA+ SRD compared to AGA-SRD. Of note, these cytokines are involved in mucosal immunity and impact homing of T cells (and other immune cells) to the colon, where gliadin can elicit a significant immune reaction (Moser and Loetscher, 2001; Ogawa et al., 2004).

Both IL-10 and IL-35 are regulatory associated immunosuppressive cytokines secreted by regulatory immune cells, including Tregs and aTregs (Wei et al., 2017). While IL-10 is broadly immunosuppressive to multiple immune cell populations, IL-35 has been shown to be more specific in suppressing T cell function. (Soltani et al., 2022; Wei et al., 2017). We found that IL-35 is increased in AGA+ SRD compared to AGA-SRD, but interestingly we did not find IL-10 to be different between AGA+ or AGA-SRD. IL-35 secreting Tregs (compared with IL-10 secreting Tregs) have been identified to be a distinct subset of effector Tregs that are better primed at suppressing auto-reactive T cell function and notably have different molecular markers of activation (Wei et al., 2017). With the antagonistic relationship between pan-T cells and negative symptoms in AGA+ SRD that we have found, it is tantalizing to speculate that Tregs may be secreting IL-35 in a protective effort against the CD8+ and non-conventional T cell populations (within the pan-T cell umbrella).

We found aTregs to be the only cell lineage to be increased in proportion in AGA+ SRD but also found it to be the only cell lineage assayed to not have any statistically significant correlations with negative symptoms. Helper T cells and Tregs notably have similarly negative correlation values in SANS Total, SANS Alogia, and SANS anhedonia categories. It may be significant as to why we do not see this correlation trend with negative symptoms continue with aTregs when it is present in both Helper T cells and Tregs. CD45RA-“aTregs” are Tregs that have encountered antigen, are T cell receptor limited, and are thus “effector” Tregs (Santegoets et al., 2015). There are several different effector Tregs – one of which may include the IL-35 secreting Tregs previously mentioned – that would fall into the larger group of Foxp3+ T cells. While we found the CD45RA-subset of Tregs to not appear to be protective against negative symptoms in AGA+ SRD, there are likely other populations of CD4+CD25+FoxP3+ T cells within the “Tregs” category (as defined in this study) that could further account for the protective effect that this group displays against negative symptoms.

Our small sample size is a pertinent limitation in this study. This small sample size with non-normally distributed data limits the statistical power in regression analysis (both the multiple interaction analysis and the logistic regression analysis), possibly obscuring significant correlations. In the case of the censored cytokine data, the simplification of converting numerical values to binary categorical values (i.e. detectable and non-detectable) does introduce a bias that likely underestimates the true difference in serum cytokine concentrations between AGA+ and negative SRD, as any value that was not “non-detectable” was converted into “detectable”. Another pertinent limitation is the limited selection of flow cytometry markers we used that may have obscured the identification of more specific T cell populations that may be involved in negative symptom pathology in AGA+ SRD. The cross-sectional nature of this study with only one time point is another notable limitation. Our findings were robust to common confounders, including age, smoking, weight, and psychotropic medication.

These findings are hypothesis-generating and illustrate several associations between T cells and negative symptoms, which is consistent with previous findings of immune dysfunction in AGA+ SRD. Our finding of an antagonizing T cell group (CD8+ and other non-conventional T cells) within “pan T-cells” that may be driving negative symptoms and a protective group of regulatory T cells within “Tregs” is particularly novel, as there is little understanding about negative symptom pathology and no specific treatments. A gluten free diet in AGA+ SRD patients has demonstrated promise as an intervention to alleviate negative symptoms in two published trials (NCT01558557 and NCT01927276) and a more recently completed replication study not yet published (NCT03183609). It will be interesting to see how a gluten free diet may affect T cell populations and the overall immune environment, as the removal of the highly immunogenic gliadin protein has already shown initial promise (Kelly et al., 2019). Further studies involving T cell function, extensive T cell subset characterization, and with data obtained at multiple time points – possibly with a gluten free diet intervention – may help to illuminate the role that T cells play in improvement of negative symptoms in this subgroup of people with SRD.

## Data Availability

All data produced in the present study are available upon reasonable request to the authors

## Supplementary Material

Supplementary Table 1 is included.

## CRediT authorship contribution statement

**Deepak Salem:** Writing – review & editing, Writing – original draft, Visualization, Formal analysis, Conceptualization. **Sarah M. Clark:** Writing – review & editing, Writing – original draft, Supervision, Methodology, Conceptualization, Data curation, Investigation. **Daniel J. Roche:** Writing – review & editing, Methodology, Conceptualization. **Nevil J. Singh:** Writing – review & editing, Methodology, Conceptualization. **Monica V. Talor:** Writing – review & editing, Methodology, Data curation. **Valerie Harrington:** Writing – review & editing. **Zhenyao Ye:** Writing – review & editing, Formal Analyses **Shuo Chen:** Writing – review & editing, Formal Analyses. **Deanna L. Kelly:** Writing – review & editing, Writing – original draft, Project administration, Conceptualization, Methodology, Data curation, Investigation.

## Acknowledgements

We acknowledge and thank the study staff and the participants whose contribution made this study possible. We would also like to thank all participating healthcare units for their cooperation during the recruitment, and the THL data collection and sample processing team. We also thank Leo Tonelli, PhD for his original assistance and the Silvio O Conte, PI and administrative team for the pilot funds to conduct the original study.

---

**Figure S1:**
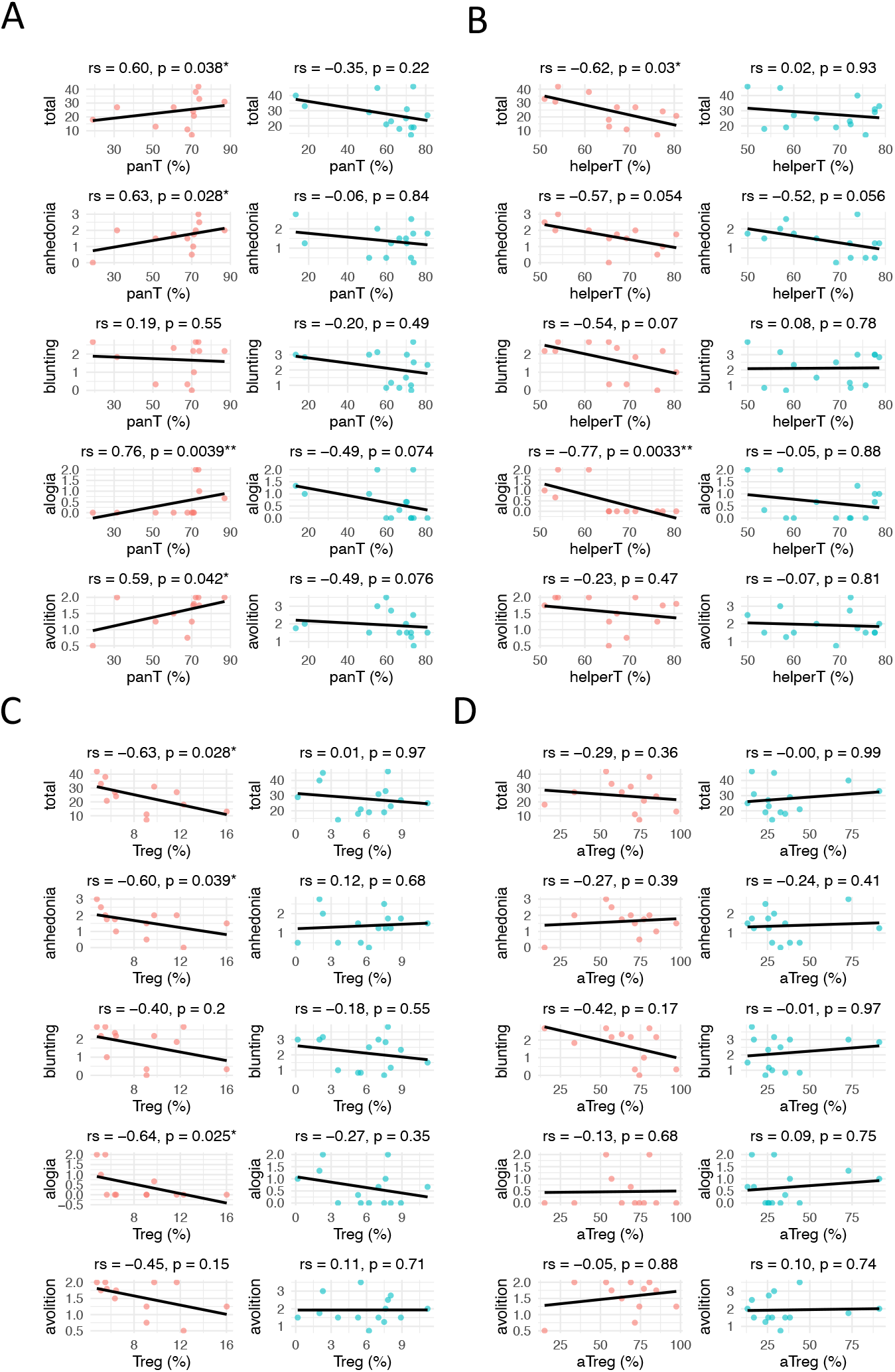
T cells correlate with negative symptoms in positive but not negative AGA-IgG schizophrenia. Spearman correlation coefficients (Spearman rs) were calculated with corresponding p values determining statistical significance. Scatterplots of each data point and a line of best fit corresponding with the correlation coefficient were generated in RStudio. A : Correlations between pan-T cells and negative symptoms. B: Correlations between helper T cells and negative symptoms. C: Correlations between Tregs and negative symptoms. D: Correlations between aTregs and negative symptoms. Abbreviations: panT (pan T cell), defined as CD3+ T cells, helperT (helper T cell), defined as CD3+CD4+ T cells, Treg defined as CD3+CD4+CD25+Foxp3+ T cells, and aTregs defined as CD3+CD4+CD25+Foxp3+CD45RA-T cells. * indicates p < 0.05; ** indicates p < 0.01.

**Table 1: Demographic information and comparisons between AGA positive and negative SRD**.

